# SARS-CoV-2 genome-based severity predictions correspond to lower qPCR values and higher viral load

**DOI:** 10.1101/2021.11.22.21266688

**Authors:** Martin Skarzynski, Erin M. McAuley, Ezekiel J. Maier, Anthony C. Fries, Jameson D. Voss, Richard R. Chapleau

**Affiliations:** Booz Allen Hamilton, Bethesda, MD 20814, USA; US Air Force School of Aerospace Medicine, Wright Patterson AFB, OH 45433, USA; US Air Force Medical Readiness Agency, Falls Church, VA 22042, USA

**Keywords:** SARS-CoV-2, COVID-19 severity, predictive modeling, genetic methods, viral epidemiology

## Abstract

The 2019 coronavirus disease (COVID-19) pandemic has demonstrated the importance of predicting, identifying, and tracking mutations throughout a pandemic event. As the COVID-19 global pandemic surpassed one year, several variants had emerged resulting in increased severity and transmissibility. In order to reduce the impact on human life, it is critical to rapidly identify which genetic variants result in increased virulence or transmission. To address the former, we evaluated if a genome-based predictive algorithm designed to predict clinical severity could predict polymerase chain reaction (PCR) results, as a surrogate for viral load and severity. Using a previously published algorithm, we compared the viral genome-based severity predictions to clinically-derived PCR-based viral load of 716 viral genomes. For those samples predicted to be “severe” (predicted severity score > 0.5), we observed an average cycle threshold (Ct) of 18.3, whereas those in in the “mild” category (severity prediction < 0.5) had an average Ct of 20.4 (*P* = 0.0017). We found a non-trivial correlation between predicted severity probability and cycle threshold (*r* = −0.199). Additionally, when divided into quartiles by prediction severity probability, the most probable quartile (≥75% probability) had a Ct of 16.6 (n=10) as compared to those least probable to be severe (<25%) of 21.4 (n=350) (*P* = 0.0045). Taken together, our results suggest that the severity predicted by a genome-based algorithm can be related to the metrics from the clinical diagnostic test, and that relative severity may be inferred from diagnostic values.

## Introduction

A classic model of disease causation is the epidemiologic triad of host, agent, and environment. Stated differently, the severity of an illness is based on an interplay between method of exposure (environment), pathogenicity of the organism (agent), and the host (host susceptibility and host response to the infectious agent). The recent SARS-CoV-2 pandemic has demonstrated that substantial diversity in both the host and virus can lead to a wide spectrum of clinical outcomes. Early research into symptom severity largely focused on host phenotypes, such as blood type [Goker 2020], age [O’Driscoll 2020], gender [Pradhan 2020], etc. As the scale of the pandemic grew, the role of geographic region and viral mutations in severe clinical outcomes began to emerge [Zhao 2020], followed by additional insights on host genetic susceptibility [COVID19 Host Genetics Initiative 2020]. Finally, efforts to predict a patient’s outcomes using computational models developed with phenotypic, genetic, and demographic data are being pursued in order to tailor patient care and manage resources [Voss, 2021; Dong 2020, Marin 2021].

Early identification of patients at increased risk of developing severe symptoms can help preserve life and health. Estimates of viral load upon admission have been shown to be correlated with higher mortality [Choudhuri et al, 2021; Pujadas et al, 2021]. Using real-time PCR data, Choudhuri and colleagues demonstrated that increased cycle thresholds were associated with 9% reduction in the odds of in-hospital mortality [Choudhuri et al, 2021]. The greatest difference was found for those patients reporting to the hospital with a cycle threshold below 23; these patients encountered 3.9-fold increased odds of in-hospital mortality compared to patients with cycle thresholds above 33. Overall, using PCR cycle thresholds as a predictor of outcome the area under the curve was found to be 0.68, suggesting an effective but limited discriminative ability for severity classification by cycle threshold at the time of admission.

The use of SARS-CoV-2 genome-wide sequencing has identified numerous variants that the World Health Organization has subsequently declared “variants of concern” [Mercatelli 2020]. Many of these variants are in the spike protein necessary for host recognition [Becerra-Florez 2020], but many are also scattered throughout the remainder of the viral genome [Nazario-Toole 2021]. Using data from the first year of the pandemic, we previously developed an algorithm to predict severity based upon viral mutations [Voss et al, 2021]. In addition, we reported 17 variants associated with severe clinical outcomes (OR≥2) and 67 variants associated with mild clinical outcomes (OR≤0.5). The area under the curve for our predictive algorithm was 0.91, suggesting a strong discriminative ability for classifying severe patients. Here we present results comparing the computationally predicted probability of a severe outcome to real-world laboratory measured PCR values.

## Materials and methods

This study was approved as a portion of the study FWR20190037N, reviewed and approved by the Air Force Research Laboratory’s Institutional Review Board. Genome sequences were downloaded from the public access GISAID database [Elbe, 2017; Shu, 2017] restricted to sequences uploaded by the US Air Force School of Aerospace Medicine (USAFSAM) as of 16 April 2021 (see supplemental information for a complete list of accession numbers and references). Only the USAFSAM sequences were used as we also have access to PCR cycle thresholds for those data. Clinical rt-PCR diagnostic tests were performed using US Centers for Disease Control and Prevention assay as described elsewhere [Chapleau et al, 2021] and the cycle threshold (Ct) for the N1 gene was used as the comparator.

A predictive algorithm based upon SARS-CoV-2 genomic variants, the geographic region of collection, and the age and gender of the patient has been reported to have be significantly predictive of clinical severity [Voss et al, 2021]. This algorithm was used to generate a predicted severity score from 0 to 1 in the Python environment. The genomic sequences were first downloaded and aligned to the Wuhan reference strain (NCBI: NC_045512.2; GISAID: EPI_ISL_402125) using MiniMap2 (version 2.17) [Li, 2018]. Sequences were aligned using MAFFT [Katoh 2002] and variants were called using SNP-sites [Page 2016]. Within our dataset, only 662 of the 4,502 variants used in the algorithm were observed in at least one viral genome, so the remaining variants were assigned columns of 0 within the Pandas dataframe (Figure 1).

**Fig 1:**
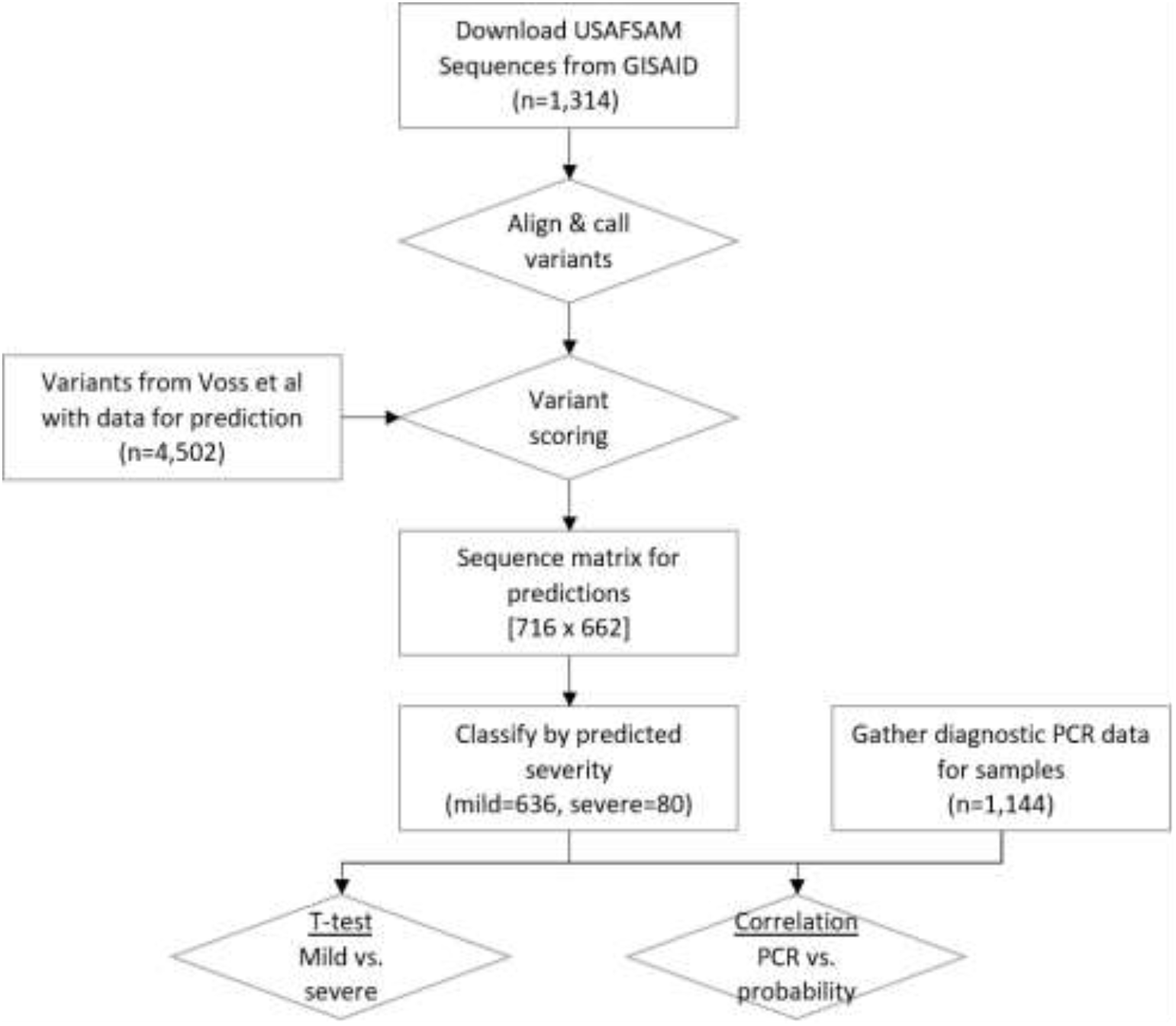
Process flow chart for orthogonal validation of a severity prediction algorithm. Sequences were downloaded from GISAID, processed through reference alignment and variant calling, predicted based upon variants identified by Voss et al, and compared to observed PCR threshold data by T-test and correlations. The matrix used for predictions was 716 samples with 662 variants overlapping the Voss et al variant list (all other variants were assigned 0 for Python predictions).

The sample IDs were divided into classes based upon severity prediction probabilities generated here for analysis. The mean observed Ct values for each class were compared using unequal variance unpaired t test. We used Welch’s corrections because of unequal sample sizes. A second approach tested for correlation between the predicted probability of a viral sequence belonging to the severe class and the observed Ct value. No modification to the original algorithm using PCR measurements was performed. Linear correlation was tested using Pearson’s correlation. We used GraphPad Prism 7.0c for performing the t tests and Pearson’s correlation.

## Results

We obtained 1,314 SARS-CoV-2 sequences published to GISAID by the USAFSAM Epidemiology Surveillance Laboratory (collection dates between 22 February 2020 and 11 June 2021). Of those sequences, 716 had corresponding N gene PCR data and were used for analysis. Using the previously published SARS-CoV-2 genome severity prediction model [Voss et al], 636 (89%) specimens had a ≤50% chance of being severe (“predicted mild” group) and 80 (11%) had a probability >50% (“predicted severe” group). The “predicted severe” group had a significantly lower observed N gene Ct value (18.3 ± 0.6) as compared to the “predicted mild” group (20.4 ± 0.2). This difference of 2.1 ± 0.7 Ct (95% CI: 0.81-3.4) is consistent with differences seen in other studies signifying highly transmissible variants of concern [Roquebert 2021] and is significant based upon the unpaired t test (Figure 2, *P* = 0.0017).

**Fig 2:**
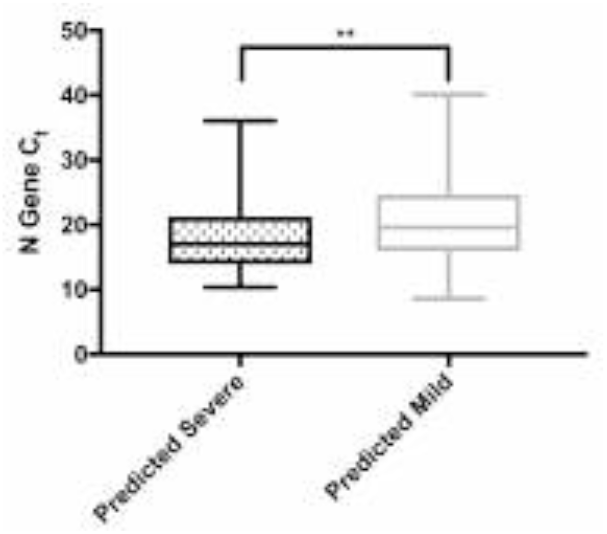
PCR Ct values for viral specimens predicted to be from patients with mild (open box, n=636) or severe (dotted box, n=80) outcomes. ^**^*P* = 0.0017.

A similar analysis on the upper and lower quartiles of severity prediction revealed a more substantial difference (Figure 3). Those specimens with the greatest probability of severe outcomes (≥75% probability) had a Ct of 16.6 ± 1.3 (n=10, 1.4% of samples), whereas those least likely to be associated with severe outcomes (<25%) had a Ct of 21.4 ± 0.3 (n=350, 49% of samples) (*P* = 0.0045). The difference between the means and 95% confidence interval was 4.9 (CI: 1.9-7.8).

**Fig 3:**
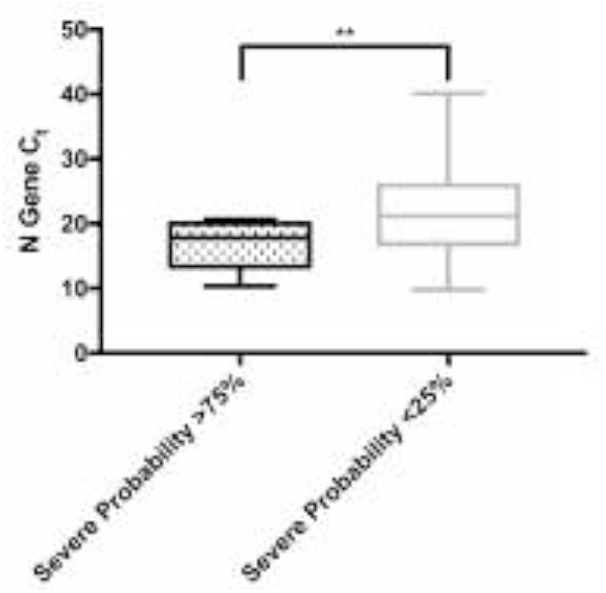
PCR Ct values for viral specimens least likely (probability <25%, open box, n=350) or most likely (probability ≥75%, dotted box, n=10) outcomes. ^**^*P* = 0.0045.

## Discussion

Thoroughly validating predictions from machine learning models is necessary for establishing the credibility and power of analytics for public health. For biology and medicine, this often means finding a testable measurement that represents the predicted outputs. In our case, we used an outgroup sample with an orthogonal severity marker to provide strong evidence that the algorithm can identify strains that are biologically unique and present meaningful clinical differences.

Research on the MERS coronavirus showed that viral load, as inferred from PCR cycle threshold, is associated with risk of severe disease and death; with a 17% higher risk of death for each 1 point drop in Ct after adjusting for age and underlying illness [Feiken, et al., 2014]. In the context of SARS-CoV-2, data from Brazil also suggests cycle threshold can predict severity in a manner that is not explained by age [Faíco-Filho et al., 2020]. Indeed, the relationship between cycle threshold and COVID-19 severity has also been identified in a prior systematic review of 18 studies with a positive association in a majority [Rao et al., 2020], while also being described in a more recent narrative review [Rabaan et al., 2021]. Typically a Ct value of less than 20 is considered “highly infective” [Englemann 2021]. Here, our results are consistent with these previous observations in that those viral genomes with the greatest predicted probability of severe clinical outcomes had, on average, a 5 Ct difference. This difference corresponds to viral titers of approximately 1.1 million viable viral particles per mL for those most likely to be severe versus only 33,000 particles per mL for those least probable to be severe.

The practical implications of Ct differences are critical to consider when determining future public health responses. n a recent study of military basic trainees, Marcus and colleagues found three interesting differences for index patients: those index patients who were symptomatic had a 7 cycle lower Ct than those who were asymptomatic (20 vs. 27.2 cycles), symptomatic index patients had a 4 cycle lower Ct than symptomatic patients who did not occur as part of a cluster (20 vs. 24.4 cycles), and those who were asymptomatic had a 9 cycle difference (27.2 for asymptomatic index patients vs. 36.4 for asymptomatic non-index patients) [Marcus 2021]. Another study found a significant difference in viral loads in a cohort of military beneficiaries associated with obesity and inpatient or outpatient status [Epsi 2021]. When inpatients and outpatients were combined, the authors did not observe any significant difference in viral load. The greatest difference observed by the authors was in severely obese outpatients, who had 77-fold higher viral loads than non-obese patients, suggesting that in mildly ill patients, those who are more obese can tolerate greater viral loads. Consequently, it becomes apparent that viral load alone is not the most powerful predictor of disease severity.

Ultimately, while it is likely that there are other factors (biological or methodological) contributing to severity, it is possible these balance out in a large enough population or where these factors are less likely to be different. For example, if there are measurement errors in the Ct associated with random differences in sample acquisition techniques, time of day a participant was tested, or sample type (nasopharyngeal vs. anterior nasal vs. nasal wash vs. oropharyngeal), these factors are likely to make the association between Ct and predicted risk appear smaller than it is. Alternatively, it remains plausible that predicted severity from the genome does not differ by time of collection or body weight category, as both the biological and methodological confounders primarily bias toward the null [Fosgate 2006]. Consequently, it is possible the difference we found is explained by factors like those mentioned above, especially considering the constrained metadata regarding the lack of patient outcomes in our dataset. Regardless, because we observed, in an independent patient sample set, a decrease in cycle thresholds (a surrogate for increased disease severity) between groups of patients predicted to experience severe clinical outcomes like hospitalization, it remains compelling that the predictions generated from the computational algorithm are reflecting biological mechanisms and are clinically meaningful for patients, providers, and public health leaders. Future work is required to take the results of these two papers from statistically significant research findings to practical tools for use in clinical practice and public health management.

In conclusion, here we report a correlation between a computationally predicted severity and a clinically measured surrogate for severity, PCR cycle threshold. Our results show that using viral genetic information combined with patient demographics could aid clinical triage and public health surveillance. While whole viral genome sequencing of every patient upon admission is cost prohibitive and results are unlikely to be timely enough to improve patient treatment, rapid diagnostic tests developed to identify variants critical to severity may provide insights to improve outcomes. Additionally, using sequence data along with laboratory validated, *in silico*-derived severity markers could help prioritize new variant vaccine development and in cases where there is clinical equipoise (for example, in cases where monoclonal antibody treatment is being considered or where post-exposure pre-symptomatic prophylaxis is being considered for known contacts, among others). Finally, public health officials employing large scale surveillance methods could benefit from these *in silico* markers by watching for the emergence of markers and gain a few days’ lead time for preparedness actions.

## Limitations

One limitation of this study is the use of PCR cycle threshold as a surrogate for clinical severity. As mentioned in the main text, the ability of PCR to predict clinical outcome only has an AUC of 0.68, making it a fair, but not ideal analog for clinical outcomes. Clinical outcome data were not available for this study. Another limitation related to using the PCR Ct value is the variability in sample collection per patient as some patients report to clinics early in illnesses whereas others may have reported only after symptoms became more severe. Regardless of these limitations, the high predictive ability of the algorithm along with the significant correlation with PCR data suggest that our results can be replicated in other populations and by other laboratories.

Another limitation is the relatively small number of viral genotypes. A post-hoc power analysis for the predicted severe (Ct mean = 18.33 ± 5.514, n=80) versus predicted mild (20.44 ± 5.633, n=636) with a 5% type I error rate revealed 89.6% power to detect the observed difference, indicating that though the sample sizes were relatively small, the study had sufficient power to draw the conclusions from the results. Similarly, the post-hoc power analysis for the most and least likely to be severe groups had a 95.3% power.

## Data Availability

All data produced in the present work are contained in the manuscript

## Declarations

### Ethics approval and consent to participate

This study was approved by the Air Force Research Laboratory’s Institutional Review Board (FWR20190037N).

## Acknowledgments

The authors thank the laboratory technicians of the USAFSAM Applied Technology and Genomics Division for their support in generating the SARS-CoV-2 genomic sequences. The authors also thank the staff of the USAFSAM Public Health Epidemiology Surveillance Laboratory for their support and coordination of sample acquisition and PCR cycle threshold data.

## Supplemental file

Excel file, two tabs. Tab 1: list of GISAID identifiers used; Tab 2: results of prediction (binary), probability of severity, and observed Ct values (if any).

